# Assessing the Complex Relationship Between Urban Tree Canopy and Mental Health in Alabama: The Role of Socioeconomic Factors

**DOI:** 10.1101/2025.11.12.25340089

**Authors:** Simona Shirley, Rajarshi Dey, Bisakha Sen, Elizabeth H. Baker, Laurie A. Malone

## Abstract

Urban tree canopy, or the proportion of city land surface covered by trees as viewed from above, has been studied as an ecological resource with various physical and mental health benefits. At the same time, disparities in both the availability and quality of trees by neighborhood disadvantage have been noted, and the relationship between tree canopy and health is sometimes ambiguous. This study employs observational, secondary data to study the associations between urban tree canopy and self-reported mental and physical health in Alabama, a state with a history of redlining that contributed to neighborhood disadvantage and environmental degradation. Multivariate linear regression models were estimated to explore the relationship between self-reported poor mental and physical health and tree canopy aggregated at the census-tract level in Alabama, USA. A stepwise modeling framework initially revealed a negative correlation between poor mental health and less extensive tree canopy. However, this relationship was suppressed when lack of insurance, which had a strong correlation with poverty, was included. It is likely that in areas of higher poverty, more extensive tree canopy may even exacerbate poor health. It seems likely that, for tree canopy to benefit health and well-being, it is important to consider the quality of the neighborhood trees and to address concerns the community may have regarding maintenance and safety issues around neighborhood trees.

## Introduction

Exposure to the natural environment is increasingly recognized as beneficial to human health, and a growing body of research has demonstrated the extensive health benefits of nature-centered interactions, including improved physical and mental well-being [1]. Recently, attention has increasingly focused on a specific and often undervalued component of nature: the tree.

Trees can offer a range of health benefits, including improving respiratory health by filtering air pollution, reducing health risks through ultraviolet radiation protection, mitigating heat-related illnesses by lowering ambient temperatures, and encouraging more physical activity in outdoor environments [1]. In urban areas, where green spaces are often replaced by concrete infrastructure, there is renewed interest in preserving and expanding the proportion of a city’s land surface covered by trees as viewed from above, also known as *tree canopy*, to improve the health of residents [2].

Associations between urban green space and mental health are also drawing attention. In North Carolina, access to and quality of green space has been linked to improved youth mental health [3]. In Australia, green space was linked to reduced social loneliness among adults [4].

Furthermore, urban tree canopy may provide superior mental health benefits compared to other forms of green space. For example, a study conducted in Singapore found that tree canopy had the strongest association with mental health compared to other measures, such as vegetation cover and park area [5]. Similarly, research in Southern California found that maternal exposure to trees was more closely associated with reduced postpartum depression than exposure to other types of green space [6]. In Jefferson County, KY, neighborhoods with less extensive tree canopy were found to have higher rates of suicide and homicide [7]. At the same time, the distribution of urban tree canopy in the United States is often unequal, and historically marginalized communities, particularly those with predominately minority populations, commonly have significantly less tree canopy than more socioeconomically advantaged communities [8].

This study evaluated the relationship between tree canopy and mental health in urban areas of Alabama, a state that consistently ranks among the lowest in the United States for several health outcomes and exhibits significant health disparities across socioeconomic groups [9]. Though the state is one of the more heavily forested in the country, substantial disparities exist in urban tree canopy [10,11]. For example, in the city of Birmingham, maps of historical redlining closely mirror current tree canopy distribution [11], with limited vegetation contributing to higher ambient temperatures in disadvantaged neighborhoods [12]. Little research exists on the relationship between urban greenspace and health in Alabama. This study helps fill this gap by utilizing publicly available health data and newly obtained tree canopy data from the state of Alabama to evaluate the relationship between urban trees and self-reported health among the adult population.

## Materials and Methods

### Methods

This ecological study examined the relationship between urban tree canopy and self-reported poor mental health across census tracts in the state of Alabama that were designated as “urban.” As a comparison, we decided to include additional models that substituted “self-reported poor mental health” with “self-reported poor physical health” as the outcome. All variables were aggregations at the census tract level and were obtained from the U.S. Climate Vulnerability Index [13], which compiles data from multiple sources into one publicly available resource. The only exception is the tree canopy data, which were obtained from the Social Determinants of Health (SDH) Core Program at the University of Alabama at Birmingham. Our final sample includes a total of 618 tracts in the analysis, which had no missing data.

Multivariate linear regression models were used to estimate the association between tree canopy and self-reported health at the census-tract level. The primary outcomes of interest were the percentages of adults reporting poor mental health and poor physical health within the past 30 days. These measures were derived from self-reported survey data, with higher percentages indicating worse health outcomes.

The main explanatory variable was the percentage of land area covered by tree canopy in each census tract, which helped capture urban greenness. In addition, to account for potential confounding, a series of covariates were introduced in a stepwise modeling framework. We decided to include separate sociodemographic, economic, and environmental covariates instead of using aggregated indexes, such as the Area Deprivation Index or Social Vulnerability Index, to better understand the association of specific census-tract-level characteristics with the outcomes of interest. This approach necessitated exploring the correlation between variables as well as multicollinearity tests to ensure that the variables selected were not highly collinear. Using this approach, the final variables chosen were as follows. Two sociodemographic factors were included—the percentage of residents aged 65 years or older and the percentage of minority population in the census tract. Two additional environmental exposures beyond tree canopy were selected—the percentage of housing units at risk for lead paint exposure and the average concentration of fine particulate matter (PM2.5) in the atmosphere. Finally, a measure of economic disadvantage was selected—the percentage of adults lacking health insurance.

Notably, this variable is highly correlated to census-tract-level poverty rate but was selected over poverty rate due to its stronger predictive ability and lower collinearity with other variables, though sensitivity analyses were conducted. Specific definitions for each variable are presented in the Data section.

Linear regression models were constructed separately for each outcome variable. Four models were estimated for both mental and physical health outcomes. Model 1 included only tree canopy coverage. Model 2 added sociodemographic factors (age 65+ years and minority percentage). Model 3 also incorporated environmental exposures (lead paint and PM2.5). Model 4 additionally introduced the percentage of uninsured adults.

Model performance was evaluated using adjusted R-squared values. Statistical significance was assessed using a 0.05 level of significance. All analyses were conducted using standard linear modeling procedures in R (version). Robustness checks with poverty rates used in place of lack of insurance were also conducted.

This study is the first to our knowledge to explore the relationship between tree canopy and mental health in Alabama. We hypothesized that self-reported poor mental health and tree canopy would demonstrate a significant negative correlation, even after adjusting for potential confounding in all four models.

### Data

#### Tree Canopy Data

Tree canopy data were obtained from the SDH Core Program at the University of Alabama at Birmingham. Tree canopy was measured at the census tract level. It was calculated using the U.S. NLCD Tree Canopy Cover produced by the Multi-Resolution Land Characteristics Consortium for the National Land Cover Database. Tree canopy coverage represents the proportion of land surface covered by trees between the years 2011 to 2021 for each pixel measured at 30-m resolution. Averages across these pixels were performed using the zonal statistics as a table tool in ArcGIS Pro; 0% indicated no trees present while 100% indicated full tree canopy coverage. The measure was developed by Liz Baker, PhD, SDH Core co-director, with help from Katie Sweeney, a PhD student in medical sociology.

#### Mental Health and Physical Health Data

Self-reported mental health and physical health data were obtained from U.S. Climate Vulnerability Index [13]. The index is a free online tool created by the collaborative efforts of the Environmental Defense Fund, Texas A&M University, and Darkhorse Analytics (Edmonton, Alberta, Canada).

Mental health data were listed as “Self-Reported Mental Health,” which was defined as the “age-adjusted prevalence of adults who report 14 or more days during the past 30 days during which their mental health was not good.” Therefore, higher percentages in the dataset indicated poorer mental health. The definition of mental health was subjective and included factors such stress, depression, and problems with emotions. The detailed probability of adults aged 18 years or older who report poor mental health for 14 or more days during the past 30 days was determined by using data from the Centers for Disease Control and Prevention’s Behavioral Risk Factor Surveillance System (2018, 2019), U. S. Census Bureau data from 2010, and American Community Survey (ACS) estimates from the years 2015–2019 or 2014–2018 to create a multilevel regression and post-stratification approach.

Physical health data were listed as “Self-Reported Physical Health,” which was defined as the “age-adjusted prevalence of adults who report 14 or more days during the past 30 days during which their physical health was not good.” Therefore, higher percentages in the dataset indicated poorer physical health. The definition of physical health was subjective and included both physical illness and injury. The detailed probability was calculated using the same model and datasets as Self-Reported Mental Health.

#### Sociodemographic, Environmental, and Economic Data

Data for minority status, aged 65 years and older, the percentage of housing units built before 1960 (lead paint risk), annual average PM2.5 concentrations, and current lack of health insurance were obtained from the U.S. Climate Vulnerability Index [13]. The U.S. Climate Vulnerability Index obtained data on minority status from the Centers for Disease Control and Prevention’s Social Vulnerability Index, which is based on results from the 5-year ACS and was released in 2018. Data representing minority status is based on percentile ranking, includes all ethnic and racial minorities, and were collected at the census tract level. The percentile ranking of populations aged 65 years or older from the U.S. Climate Vulnerability Index was also derived from the 2018 ACS data collection at the census tract level.

The environmental factor accounting for the percentage of housing units at risk for lead paint exposure was listed as “Lead Paint: % housing units built before 1960” in the U.S. Climate Vulnerability Index. Percentages were obtained from the U. S. Environmental Protection Agency and based on 2020 U.S. Census Tract data. Data for annual average PM2.5 concentrations were listed at the census tract level as a 3-year average between 2017 and 2019. The data for annual average PM2.5 concentrations was obtained from a 2021 study published in the peer-reviewed scientific journal *Environmental Science & Technology* [14].

The economic factor “current lack of health insurance” was defined by the U.S. Climate Vulnerability Index as the “age-adjusted prevalence of adults aged 18 to 64 years who report having no current health insurance” at the census tract level. This variable was originally derived as a detailed probability using 2018–2019 data from the Behavioral Risk Factor Surveillance System, 2010 U.S. Census Bureau data, and ACS estimates from 2015–2019 or 2014–2018.

## Results

### Analysis of Variables

Detailed descriptive statistics for all variables are presented in Table 1. The mean prevalence of self-reported poor mental health was 67.2% (95% CI: 65.0–69.4), while the mean for poor physical health was 58.6% (95% CI: 56.1–61.0). Tree canopy averaged 36.5% (95% CI: 35.2–37.8), with substantial variability across census tracts. Other covariates, including minority percentage, age 65 years and older, and lack of insurance, also showed wide distributions.

**Table 1.**
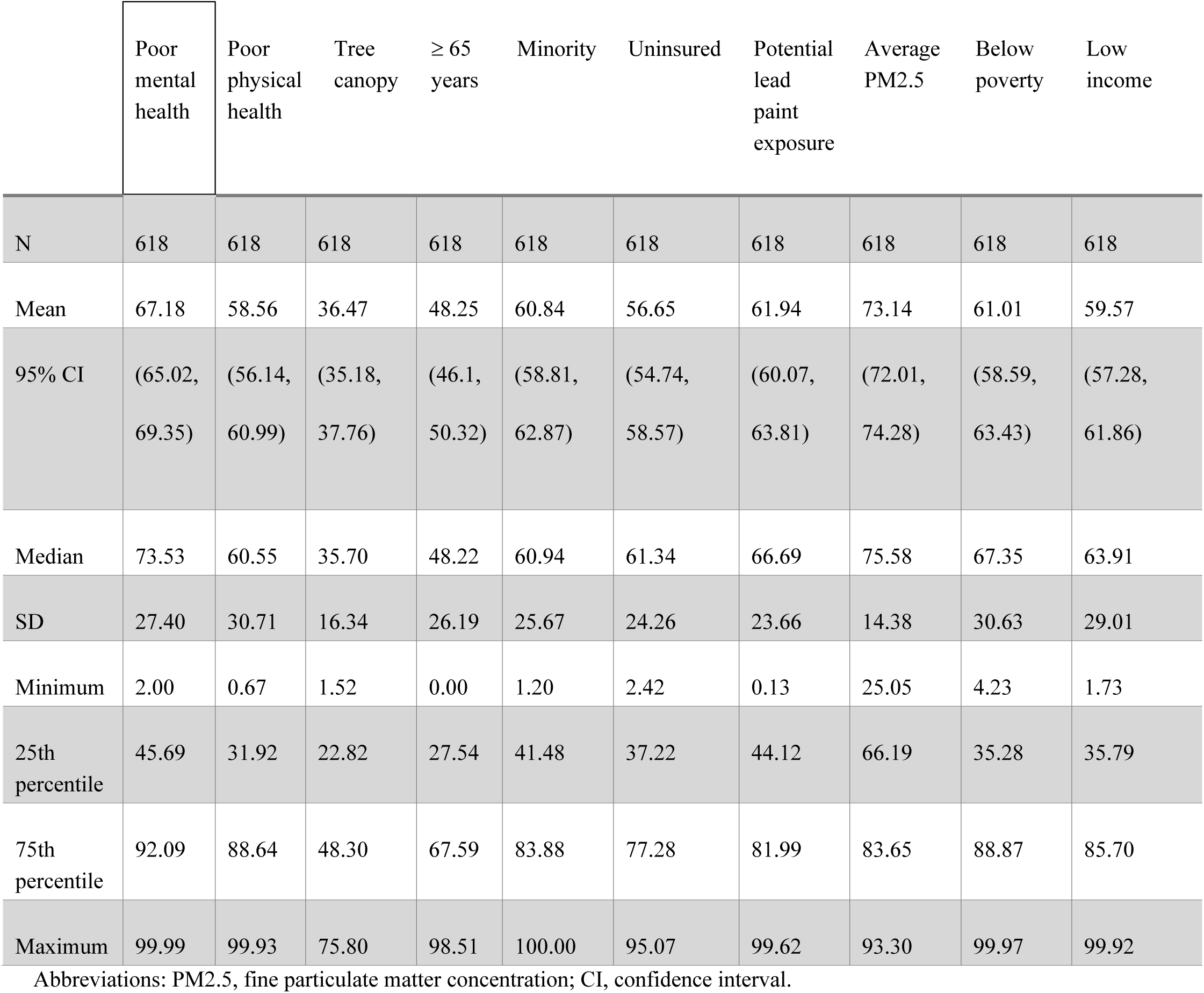
Descriptive statistics of variables considered

|  | Poor<br>mental<br>health | Poor<br>physical<br>health | Tree<br>canopy | ≥ 65<br>years | Minority | Uninsured | Potential<br>lead<br>paint<br>exposure | Average<br>PM2.5 | Below<br>poverty | Low<br>income |
| --- | --- | --- | --- | --- | --- | --- | --- | --- | --- | --- |
| N | 618 | 618 | 618 | 618 | 618 | 618 | 618 | 618 | 618 | 618 |
| Mean | 67.18 | 58.56 | 36.47 | 48.25 | 60.84 | 56.65 | 61.94 | 73.14 | 61.01 | 59.57 |
| 95% CI | (65.02,<br>69.35) | (56.14,<br>60.99) | (35.18,<br>37.76) | (46.1,<br>50.32) | (58.81,<br>62.87) | (54.74,<br>58.57) | (60.07,<br>63.81) | (72.01,<br>74.28) | (58.59,<br>63.43) | (57.28,<br>61.86) |
| Median | 73.53 | 60.55 | 35.70 | 48.22 | 60.94 | 61.34 | 66.69 | 75.58 | 67.35 | 63.91 |
| SD | 27.40 | 30.71 | 16.34 | 26.19 | 25.67 | 24.26 | 23.66 | 14.38 | 30.63 | 29.01 |
| Minimum | 2.00 | 0.67 | 1.52 | 0.00 | 1.20 | 2.42 | 0.13 | 25.05 | 4.23 | 1.73 |
| 25th<br>percentile | 45.69 | 31.92 | 22.82 | 27.54 | 41.48 | 37.22 | 44.12 | 66.19 | 35.28 | 35.79 |
| 75th<br>percentile | 92.09 | 88.64 | 48.30 | 67.59 | 83.88 | 77.28 | 81.99 | 83.65 | 88.87 | 85.70 |
| Maximum | 99.99 | 99.93 | 75.80 | 98.51 | 100.00 | 95.07 | 99.62 | 93.30 | 99.97 | 99.92 |

Abbreviations: PM2.5, fine particulate matter concentration; CI, confidence interval.

### Association Between Tree Canopy Coverage and Self-Reported Poor Mental Health

Results for linear regression models where the outcome was mental health are presented in Table 2, and the four models described previously are denoted as Models A1 to A4. More tree canopy coverage was significantly associated with lower levels of poor mental health in Model A1 (β = -0.299, 95% CI: -0.37 to -0.23, p < 0.001), though this simple model explained only 10.3% of the variance in the outcome variable. In Model A2, after adjusting for sociodemographic factors, more extensive tree canopy coverage appeared to remain a significant protective factor (β = -0.091, p = 0.002). In Model A3, which added environmental exposures, the association persisted (β = -0.087, p < 0.001). However, in Model A4, the inclusion of lack of insurance reversed the direction of the tree canopy coefficient (β = 0.019, p = 0.07) and, by a small margin, it was no longer considered statistically significant.

**Table 2.**
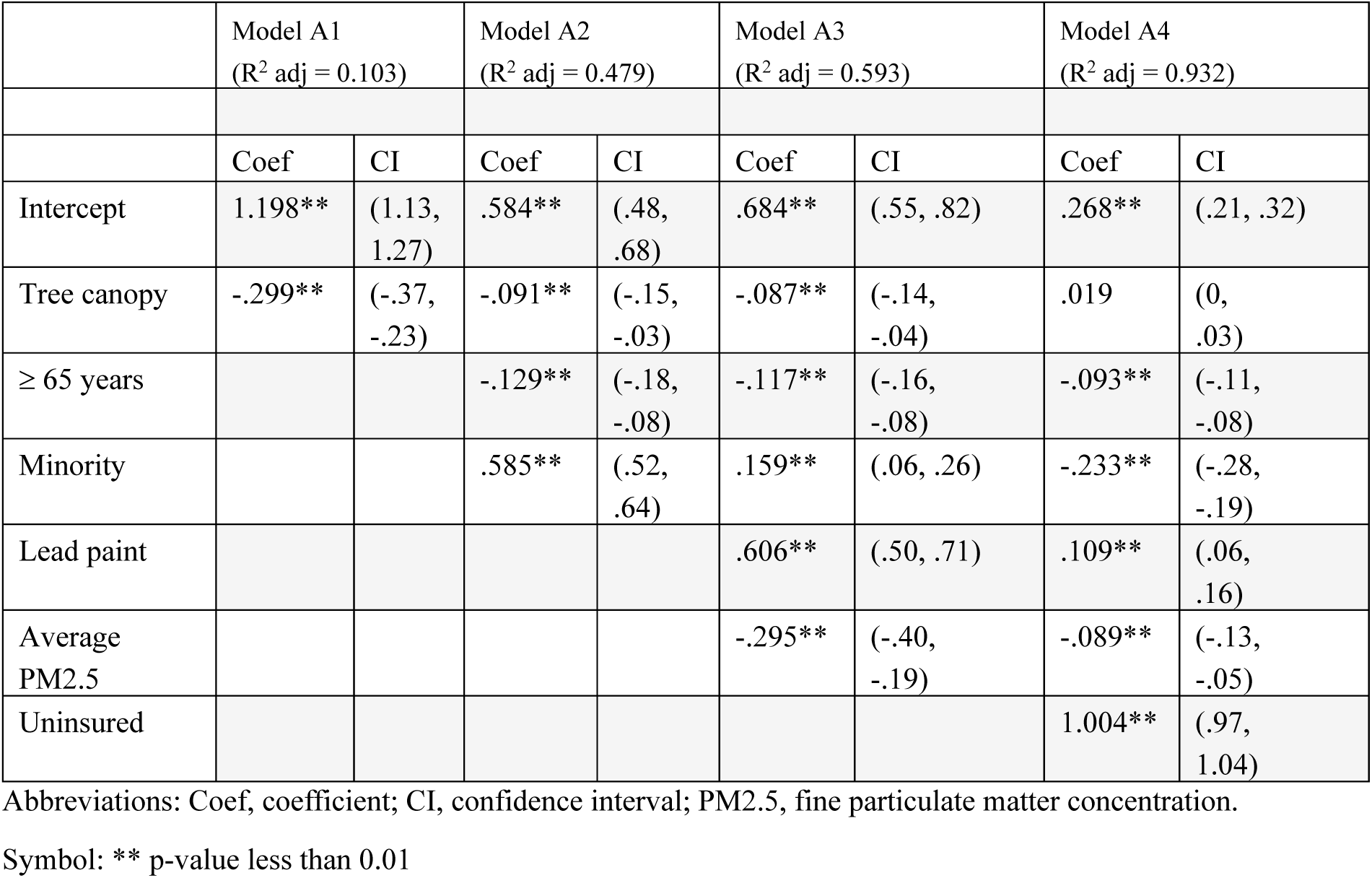
Comparison of regression models for self-reported poor mental health

The other covariates also played important roles in predicting mental health outcomes. In Model A2, both age 65 years and older (β = -0.129, p < 0.001) and minority percentage (β = 0.585, p < 0.001) were significant, suggesting that older populations were associated with lower mental stress than younger populations, while higher minority percentages were associated with higher stress. In Model A3, lead paint exposure (β = 0.606, p < 0.001) and PM2.5 (β = -0.295, p < 0.001) were significant, though the negative PM2.5 coefficient is counterintuitive and may reflect complex spatial or other confounding. Model A4 revealed that lack of insurance was a strong predictor of poor mental health (β = 1.004, p < 0.001).

### Association Between Tree Canopy Coverage and Self-Reported Poor Physical Health

Results for linear regression models where the outcome is physical health are shown in Table 3, and the models are denoted as Models B1 to B4. Tree canopy coverage was initially associated with lower levels of poor physical health in Model B1 (β = -0.289, 95% CI: -0.38 to -0.20, p < 0.001). In Model B2, after adjusting for sociodemographic factors, the tree canopy coefficient maintained a negative association with physical health but was no longer statistically significant (β = -0.037, p = 0.27). This pattern continued in Model B3, which added environmental exposures (β = -0.032, p = 0.29). However, in Model B4, the inclusion of lack of insurance reversed the direction of the tree canopy coefficient, which became significant and positive (β = 0.085, p < 0.001).

**Table 3.** Comparison of regression models for self-reported poor physical health

|  | *Model B1*<br>(*R*<sup>2</sup> *adj* = 0.063) |  | *Model B2*<br>(*R*<sup>2</sup> *adj* = 0.510) |  | *Model B3*<br>(*R*<sup>2</sup> *adj* = 0.613) |  | *Model B4*<br>(*R*<sup>2</sup> *adj* = 0.887) |  |
| --- | --- | --- | --- | --- | --- | --- | --- | --- |
|  | Coef | CI | Coef | CI | Coef | CI | Coef | CI |
| *Intercept* | 1.149** | (1.06, 1.24) | -.068 | (-.19, 0.05) | .006 | (-.15, .17) | -.453** | (.21, .32) |
| Tree canopy (%) | -.289** | (-.38, -.20) | -.037 | (-.10, 0.02) | -.032 | (-.09, .03) | .085** | (.05, .12) |
| ≥ 65 years |  |  | .231** | (0.18, 0.29) | .247** | (.20, .30) | .272** | (0.25, 0.30) |
| Minority (%) |  |  | .851** | (0.78, 0.92) | .329** | (.20, .45) | -.104** | (-.17, -.03) |
| Lead paint (%) |  |  |  |  | .731** | (.60, .86) | .183** | (.11, .26) |
| Average PM2.5 (%) |  |  |  |  | -.300** | (-.43, -.17) | -.073* | (-.14, -.01) |
| Uninsured (%) |  |  |  |  |  |  | 1.106** | (1.05, 1.16) |

Abbreviations: Coef, coefficient; CI, confidence interval; PM2.5, fine particulate matter concentration.
Symbol: \*\* p-value less than 0.01
Symbol: \* p-value less than 0.05

For both mental health and physical health, including lack of insurance reversed the direction of the tree canopy coefficient, and in the mental health model, tree canopy was no longer statistically significant. The coefficient’s sign change in this model suggests a suppressor effect of lack of insurance (which may also be considered a proxy measure for poverty) on the apparent benefits of tree canopy. In other words, in Models 1 through 3, tree canopy coverage may essentially pick up the protective effects independently provided by living in areas with greater insurance coverage and affluence. Once greater insurance coverage is accounted for, there is even the suggestion that more extensive tree canopy coverage may be associated with poorer self-reported health.

## Discussion

This ecological study explored the relationship between urban tree canopy and self-reported poor mental health across census tracts in urban areas of Alabama. Publicly available data from the U.S. Climate Vulnerability Index was combined with new tree canopy data built by the SDH Core at the University of Alabama at Birmingham. Arguably, leveraging publicly available data will become increasingly important for environmental justice research, especially for junior scholars, if funding support for such research is diminished.

The expectation that greenspace improves health is grounded in several well-established mechanisms. Trees and other forms of urban vegetation can mitigate urban heat, improve air quality, and encourage physical activity [1]. While a growing body of literature has found associations between exposure to greenspace and better health, it is still unclear to what extent underlying confounders may have influenced the results.

Our findings suggest a nuanced and context-dependent association between urban tree canopy and health outcomes. In unadjusted models, more extensive tree canopy was significantly associated with a lower prevalence of self-reported poor mental health. This finding aligns with the literature linking greenspace to improved well-being [4]. This association continued in models that adjusted for sociodemographic and environmental factors. However, after adjusting for lack of insurance, which serves as a proxy for poverty, the association between less extensive tree canopy and poor mental health reversed direction and became positive, while just falling short of statistical significance. Our study suggests that the “beneficial” association of tree canopy is driven by the underlying census tract level of economic affluence. Other works have suggested that the benefits of greenspace exposure are linked to socioeconomic affluence [15].

Thus, our findings add to the emerging literature that emphasizes the idea of a context-dependent relationship between tree canopy and mental health. In disadvantaged communities, greenspaces may be of lower quality; they may be poorly maintained, overgrown and potentially unsafe—factors that may transform a potentially valuable neighborhood resource into a disamenity [16]. For example, overgrown vegetation in vacant lots or around abandoned homes may count as “tree canopy” but will likely have deleterious psychological effects for neighborhood residents [16]. For example, in Milwaukee, WI, denser canopy in African American neighborhoods was largely due to a profusion of unmanaged “volunteer” species that emerged due to neglect [17]. In other large urban centers, minority residents have expressed resistance to urban trees due to concerns about hazards, including root damage, falling branches, pollen allergies, and maintenance responsibilities [18,19].

These findings underscore the need to move beyond simplistic metrics of greenness.

More extensive tree canopy alone is not a panacea for health improvement when economic disadvantages are considered. Urban greening strategies may only prove beneficial if coupled with parallel efforts to address derelict conditions of the built environment, such as repurposing vacant lots [20]. Active community engagement in the greening process may be key for disadvantaged communities to reap the health benefits associated with urban trees and to avoid the detrimental effects of green gentrification [20–22].

This study has several strengths, including the use of a large, population-based dataset, a stepwise modeling approach that allows for disentangling of complex relationships, and the inclusion of various confounding variables from sociodemographic, environmental, and economic outlooks. The use of novel tree canopy data from Alabama provides a rare perspective from the Southeastern United States, a region rarely included in discussions about greenspace and health.

We acknowledge several limitations. The observational design precludes causal inference, and the reliance on self-reported health measures may introduce reporting bias. Additionally, the cross-sectional nature of the data limits our ability to assess temporal relationships. Census-tract-level aggregations prevent more nuanced analyses at the individual level. For example, recent research suggests that reductions in ambient temperature and any resultant benefits are most dependent on proximity and density of tree cover within 10 m [23], and census-tract-level measures cannot capture such variations. Tree canopy was measured via satellite imagery, which does not capture street-level quality or usability. Additionally, we could not assess species composition, maintenance status, or residents’ perceptions of greenspace, factors that may be critical drivers of the health benefits of urban trees.

Future directions include collaboration with local organizations in Alabama such as Cool Green Trees [24], which currently works to plant more trees in Alabama’s urban heat islands.

Tree canopy studies, as well as other nature-centered health studies, should continue to involve previously overlooked areas of the United States such as Alabama and the rest of the Southeast region.

## Conclusions

Despite these above-mentioned limitations, this study contributes to a growing understanding of the complex, context-dependent relationship between urban greenness and health by leveraging publicly available data and novel tree canopy estimates from Alabama, a state that has largely been overlooked in this area of research.

Ultimately, it may require large randomized controlled trials, such as the Green Heart Project in Louisville, KY [2], to rigorously ascertain the health benefits of planting urban trees. However, as funding for environmental justice research becomes more constrained, leveraging open-access datasets will be important for researchers seeking to inform equity-focused urban planning. To fully understand how urban greening strategies can provide equitable health benefits to all residents, our findings emphasize the importance of moving beyond simplistic measures of greenness and tree canopy to more comprehensively assessing the roles of community blight, safety, proper management of trees and other green spaces, and community involvement in greenspace development.

## Institutional Review Board Statement

Not applicable.

## Data Availability Statement

The original data presented in the study are openly available at climatevulnerabilityindex.org. The tree canopy data presented in this study are available on request from the corresponding author.

## Author Contributions

Conceptualization, S.S., B.S., and L.M.; methodology, B.S and R.D.; software, R.D.; validation, R.D..; formal analysis, B.S. and R.D.; investigation, S.S., L.M., B.S., and R.D.; resources, L.B. and R.D.; data curation, L.B., S.S., and L.M.; writing—original draft preparation, S.S..; writing—review and editing, S.S., L.M., and B.S.; visualization, R. D..; supervision, B.S. and L.M..; project administration, B.S. and L.M.. All authors have read and agreed to the published version of the manuscript.

## Acknowledgments

We thank Emily Delzell for her editorial review and assistance in the preparation of this manuscript.

